# Psychosocial Profiles of Older Adults by Dentition Status and Dental Utilization History

**DOI:** 10.1101/2024.05.05.24306865

**Authors:** T.L. Finlayson, K. Moss, J.A. Jones, J.S. Preisser, J.A. Weintraub

**Author notes:** Corresponding Author; Twitter: @FinlaysonTracy.

## Abstract

**Objective:** Psychosocial factors can affect health. Patterns of psychosocial stressors and resources among older adults were examined for oral health status.

**Methods:** The Health and Retirement Study (HRS) is a representative sample of US adults >50 years. Participants completed the 2018 HRS CORE survey and Psychosocial and Lifestyle Questionnaire–Panel A “Leave Behind” survey (HRS-LB)(N=4703). All measures were self-reported, and stratified into outcome groups: 1) edentulous/dentate, 2) with/without a recent dental visit in the last two years. Psychosocial measures covered three domains: well-being, beliefs, and lifestyle. Specifically, we studied loneliness, life satisfaction, perceived age, social status, control, mastery, and chronic stressors. Latent class analysis (LCA) identified profiles of adults based on the distribution of psychological and social stressors and resources. Associations between latent classes and being edentulous and a recent dental visit were examined in logistic regression models.

**Results:** About 30% reported no recent dental visit; 14% were edentulous. Three latent classes were identified; profiles had different distributions of psychosocial factors. Half were in Class A:“Satisfied/Connected” (n=2230), 27% in Class B:“Satisfied/Lonely” (n=1293), and 25% in Class C:“Unsatisfied/Lonely” (n=1180). “Satisfied/Connected” adults had the fewest psychosocial risk factors, most resources, were dentate, with a recent dental visit. “Unsatisfied/Lonely” adults exhibited the most psychosocial risk factors and fewest resources, more were edentulous and lacked a recent dental visit. “Satisfied/Lonely” adults exhibited characteristics between Classes A and C. In fully adjusted regression models, Class B adults had 1.29 (1.03-1.61) times greater odds than Class A to be edentulous, and 1.27 (1.07-1.50) times greater odds to not have a recent dental visit. Class C adults had 1.20 (0.96-1.51) times greater odds than Class A to be edentulous, and 1.33 (1.11-1.58) times greater odds to not have recent dental visit.

**Conclusion:** Adverse psychosocial factors are associated with edentulism and lack routine dental visits.

## INTRODUCTION

Social conditions can adversely affect health, but social deprivation and other stressors do not universally lead to poor health. Adults encounter various life transitions and stressors as they age, which can affect health in different ways.^1^ While individual psychological appraisals and responses to stressors may help buffer potential negative impact, there is a lack of agreement about chronic stress measurement or processes.^1^ Psychosocial factors refer to social conditions and related individual psychological responses. Most psychosocial research focuses on acute and chronic stressors and their negative impact; few studies explore resources that contribute to resilience. Evidence of the importance of accounting for social contexts and psychosocial responses to stressors and resources from longitudinal oral health cohort studies exist in other countries.^2,3^

Changes in psychological well-being are connected to the aging process, and can affect health status.^4^ Major life transitions occur with aging, including retirement, which may include loss of employer-based dental coverage, and other social status changes that can affect an older adult’s ability to seek dental care. Many older adults, especially racial/ethnic minorities, report experiencing barriers (e.g., cost, mistrust, complex medical conditions) to accessing dental care.^5^ Traditional Medicare excludes dental services, except in few emergency or medically necessary situations. Medicare-eligible adults can purchase Medicare Advantage plans, which may include dental coverage for additional premiums and co-pays; however, benefits vary widely and are not comprehensive. Adult dental benefits are optional for state Medicaid programs; most states have limited dental benefits for adults, emergency coverage only, or none at all.^6^

The US Health and Retirement Study (HRS) is an ongoing, longitudinal and nationally representative dataset on older adults’ health status, functional abilities, life transitions and social factors. Analyses of past HRS data indicate that low-income adults were less likely to have dental insurance or visit the dentist, and were more likely to experience tooth loss.^7^ Social gradients have also been documented for dental utilization in 2008 HRS.^8^ Dental users tend to be female, healthier, wealthier, dentate, and married.^9^ The role of psychosocial factors and older adults’ oral health status and dental utilization have not been explored.

The HRS is a comprehensive data source about older adults’ lives during important transitions. However, the standard biennial CORE survey has limited psychosocial data; additional data are collected in the HRS Psychosocial and Lifestyle Questionnaire (called the “Leave Behind” or HRS-LB survey, self-administered and returned by mail). Yet, HRS-LB is underutilized; in a 2018 review of HRS publications, examination of psychosocial stressors accounted for only 5% of studies.^10^ Thus, there is a unique opportunity to leverage existing national data to explore in-depth how psychological and social risk and resource factors are associated with oral health. This study links HRS-LB psychosocial measures to examine lack of recent dental utilization and total tooth loss in the HRS CORE. Focusing on measures from three domains (well-being, beliefs, and lifestyle), our objectives were to identify latent class profiles that could be easily determined clinically based on these psychosocial factors for groups of older adults with and without any natural teeth (edentulous), and with and without a recent dental visit.

## METHODS

Secondary data analyses were conducted using publicly available data from the HRS, conducted by the University of Michigan. Approximately 20,000 adults over age 50 participate over time, with additional cohorts periodically recruited.^11,12^ HRS CORE data were collected using face-to-face and telephone interviews. This cross-sectional study assesses psychosocial factors, dental utilization and edentulism.

The HRS-LB includes several in-depth validated scales across six broad domains: overall well-being, lifestyle, work, social relations/support, personality traits, and self-related beliefs.^13^ It was administered to two subsamples from the enhanced face-to-face interviews, every four years, from approximately one-half of HRS CORE participants. The analyses used de-identified 2018 HRS CORE demographic and select health variables merged with HRS-LB Subsample A select psychosocial variables of interest in three domains to create the final analytic sample (N=4703).

Two HRS CORE oral health outcome variables captured edentulism and lack of recent dental care. Respondents who answered “Yes” to “Have you lost all of your upper and lower natural permanent teeth” were considered edentulous. Respondents who answered “No” to “In the last 2 years, have you seen a dentist for dental care, including dentures?” were categorized as <u>not</u> having a recent dental visit.

HRS-LB psychosocial measures of interest covered three broad domains: well-being, beliefs, and lifestyle. HRS-LB detailed scoring instructions for each validated scale were followed to tabulate summary scale scores (averages, unless otherwise specified).^13^ Some scale scoring allowed for individual items to be missing and still produce a valid scale score. Scales were scored for assessing distributions, and component items were dichotomized for the latent class analysis (LCA).

The “well-being domain” HRS-LB measures included three validated scales reflecting loneliness, subjective well-being/life satisfaction and domain-specific well-being.

The 11-item loneliness measure^14,15^ assessed how much of the time individuals experience loneliness (response options: often, sometimes, hardly ever/never). Four items were reversed scored, with higher scores reflect more frequent loneliness. Component items were dichotomized as often/sometimes vs. hardly ever/never.

The 5-item subjective well-being/life satisfaction scale assessed agreement with statements like “in most ways my life is close to ideal” (1=strongly disagree, 7=strongly agree); lower scores reflect lower satisfaction.^16^ Component items were dichotomized as disagree vs. neither agree or disagree/agree.

The 7-item life-situation specific satisfaction scale assessed health, family life, financials and living situation (1=completely satisfied, 5= not at all satisfied); lower scores reflect lower satisfaction ^17^. Component items were dichotomized (completely/very satisfied vs. everything else.

The five constructs within the “beliefs domain” include: perceived age, perceived social status, change in status, control and mastery, and self-efficacy. Perceived age was a single question: “Many people feel older or younger than they actually are. What age do you feel?” A new variable was created for whether or not a participant felt older than his/her actual age. Participants also responded to the 8-item satisfaction with aging scale (1=strongly disagree, 6=strongly agree), where lower scores reflect feeling negatively about aging.^18,19^ Four items were reversed. Component items were dichotomized as disagree vs. agree.

The 2-item subjective social status asked participants where they saw themselves on a 10-step ladder (higher is better position) and if they perceived their status moved up, down or did not change in the last two years.^20,21^

The 10-item sense of control has 5 items related to constraints, and 5 items related to mastery (1=strongly disagree, 6=strongly agree).^22^ Higher scores reflect more constraints. Mastery is a psychosocial resource, and higher scores are better. Component items were dichotomized as disagree vs. agree.

Three separate domain-specific control/self-efficacy questions asked about extent of control over one’s health, social life, and financial situation (0=no control, 10=very much in control)^23^. Self-efficacy is a psychosocial resource, and higher scores are better and reflect greater control. Component items were dichotomized (<7 vs. 7+).

The “lifestyle domain” captured exposure to and impact of eight different ongoing chronic stressors.^24^ Participants indicated if a stressor was currently happening, lasting over 12 months, and if so, the degree to which it was upsetting (not, somewhat, or very upsetting). Recodes reflected if a stressor was happening and somewhat/very upsetting.

HRS CORE demographic variables reflected participant race/ethnicity, sex, educational attainment, marital status, household net wealth, Medicaid participation and urban residency. HRS birth cohorts were defined based on specific years of birth. Persons born before 1924 are in the Asset and Health Dynamics Among the Oldest Old (AHEAD) cohort; persons born between 1924-30 are Children of the Depression (CODA); persons born between 1931-41 are the original HRS cohort; persons born between 1942-47 are War Babies; persons born between 1948-53, 1954-59, and 1960-65 are Early, Middle and Late Baby Boomers. HRS Baby Boomer birth cohorts were combined given their similar characteristics, as were the AHEAD and CODA cohorts due to smaller sample sizes. Four birth cohort groups were used for these analyses: 1) AHEAD and CODA; 2) HRS; 3) War Babies; and 4) Baby Boomers. Additionally, three HRS CORE health variables relevant for oral health were included: current smoker, current drinker, and diabetes.

The original distributions of all variables and psychosocial scales were tabulated, and extent of missingness assessed; the final analytic dataset had no missingness across psychosocial characteristics and outcomes of interest. Descriptive statistics (means, standard deviations, Pearson correlations for continuous scaled variables; frequencies for categorical variables) were calculated for demographic and psychosocial characteristics. Some response categories for the individual psychosocial variables had small cell sizes, thus each variable was dichotomized before being entered into the latent class model (See **Supplement 1).**

Models with two-nine LCA classes were specified and compared for model fit, considering best practices in building/selecting the final model.^25^ **Table 1** shows differences in log-likelihood for two-seven class models. In the six-class model and beyond, there were classes with ≤5% of the sample. The biggest difference in log-likelihood occurred between two to three-classes. While the five-class model appeared to offer the best statistical fit, the three-class model exhibited similar patterns and was selected since it was simpler and for ease of interpretation. The three-class model also yielded the best fit based on data distributions using heatmaps (**Figure 1**).

**FIGURE 1.**
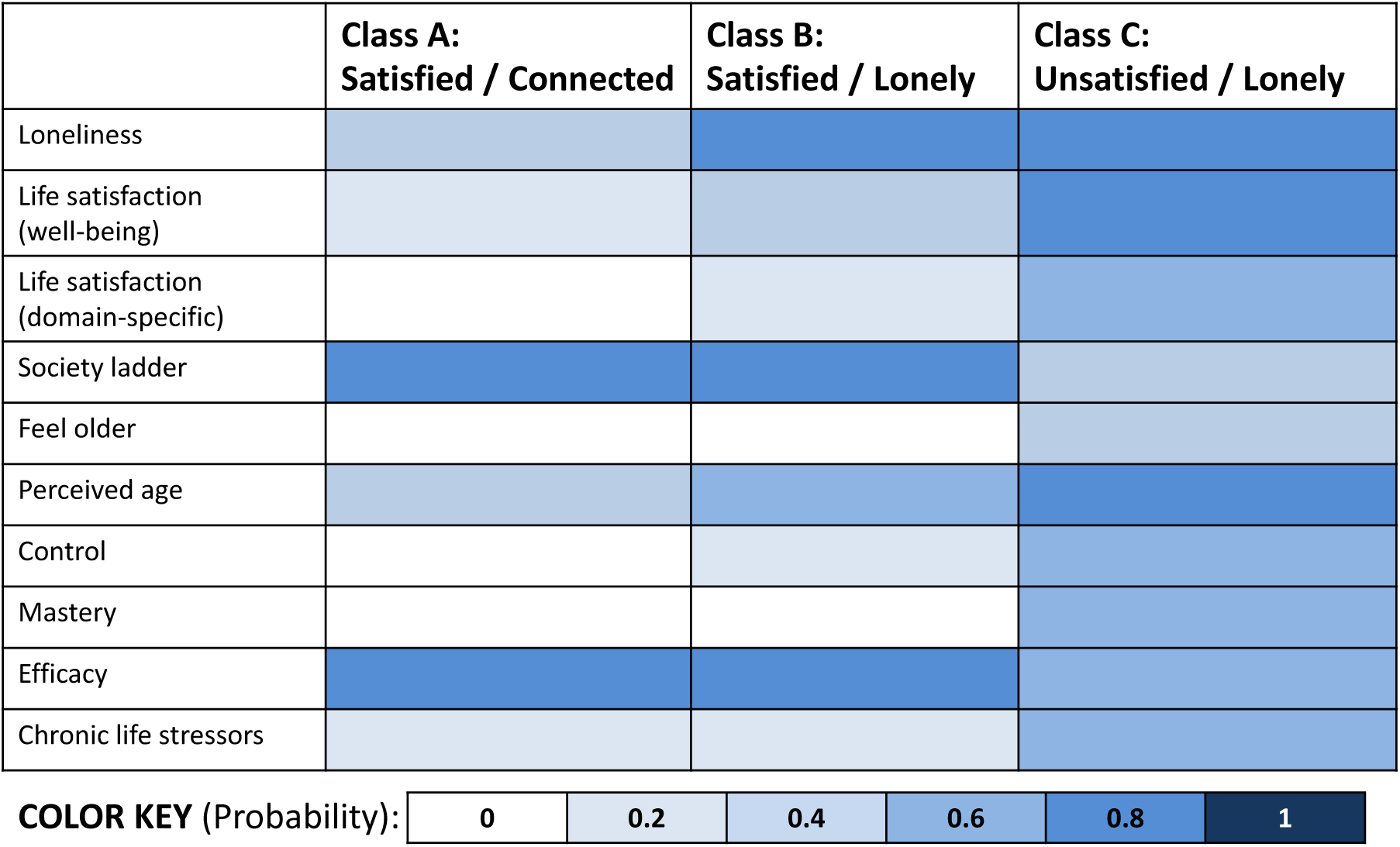
Heatmap of Latent Class Analysis (LCA) of psychosocial characteristics among older adults in the US (n=4703) Note. Data derived from LCA of sample of participants in both the 2018 Health and Retirement Study (HRS) and 2018 Leave-Back Subsample A Survey (HRS-LB). Participants were assigned to a specific class based on their posterior class membership probabilities. The color gradient shows the probability of a given characteristic conditional on class membership (darker color = higher probability).

**Table 1:** Comparison of two-seven class Latent Class Analysis Models (n=4703)

|  | 2-class | 3-class | 4-class | 5-class | 6-class | 7-class |
| --- | --- | --- | --- | --- | --- | --- |
| Log-likelihood: | -115653.9 | -112269.9 | -109808.7 | -108847.9 | -108003.1 | -107257.0 |
| Difference |  | 3384.0 | 2461.2 | 960.8 | 844.8 | 746.2 |

Study participants were categorized into the class for which they had the highest posterior probability of membership, primarily characterized by differences across self-reported levels of satisfaction with many aspects of their life (Satisfaction), and degree of social engagement and connectedness or perceptions of loneliness). Class A was characterized as “Satisfied with Life and Socially Engaged and Connected” (“Satisfied/Connected”); Class B was “Satisfied with Life, but Lonely” (“Satisfied/Lonely”), and Class C was “Unsatisfied with Life and Lonely” (“Unsatisfied/Lonely”). Each Class represented profiles of lower, moderate, or higher psychosocial risk characteristics, respectively. Descriptive statistics for demographic and psychosocial factors were calculated for each class for the full HRS-LB sample. Logistic regression models were fitted to estimate the association of the latent classes, using Class A as the reference group, with no recent dental visit in the last two years and edentulism outcomes in the HRS-LB sample (n=4703).

Minimally and fully adjusted regression models were fitted. Minimally adjusted models included race, sex, and birth cohort in addition to the latent risk class. Fully adjusted models additionally adjusted for education, marital status, wealth, Medicaid, urban, tobacco smoking, alcohol consumption, and diabetes. For dichotomous outcomes, model-adjusted odds ratios and 95% confidence intervals were computed for Classes B and C relative to Class A, with estimates exceeding 1.0 indicating an increase in risk for a worse outcome given an increase in risk level according to psychosocial latent class membership. Analyses were conducted using SAS v9.4 (SAS Institute, Cary, NC).

## RESULTS

**Table 2** summarizes the distribution of demographic and psychosocial characteristics for all participants in HRS-LB, stratified by dentition status, dentate or edentulous, and by recent dental utilization (yes/no).

**Table 2:**
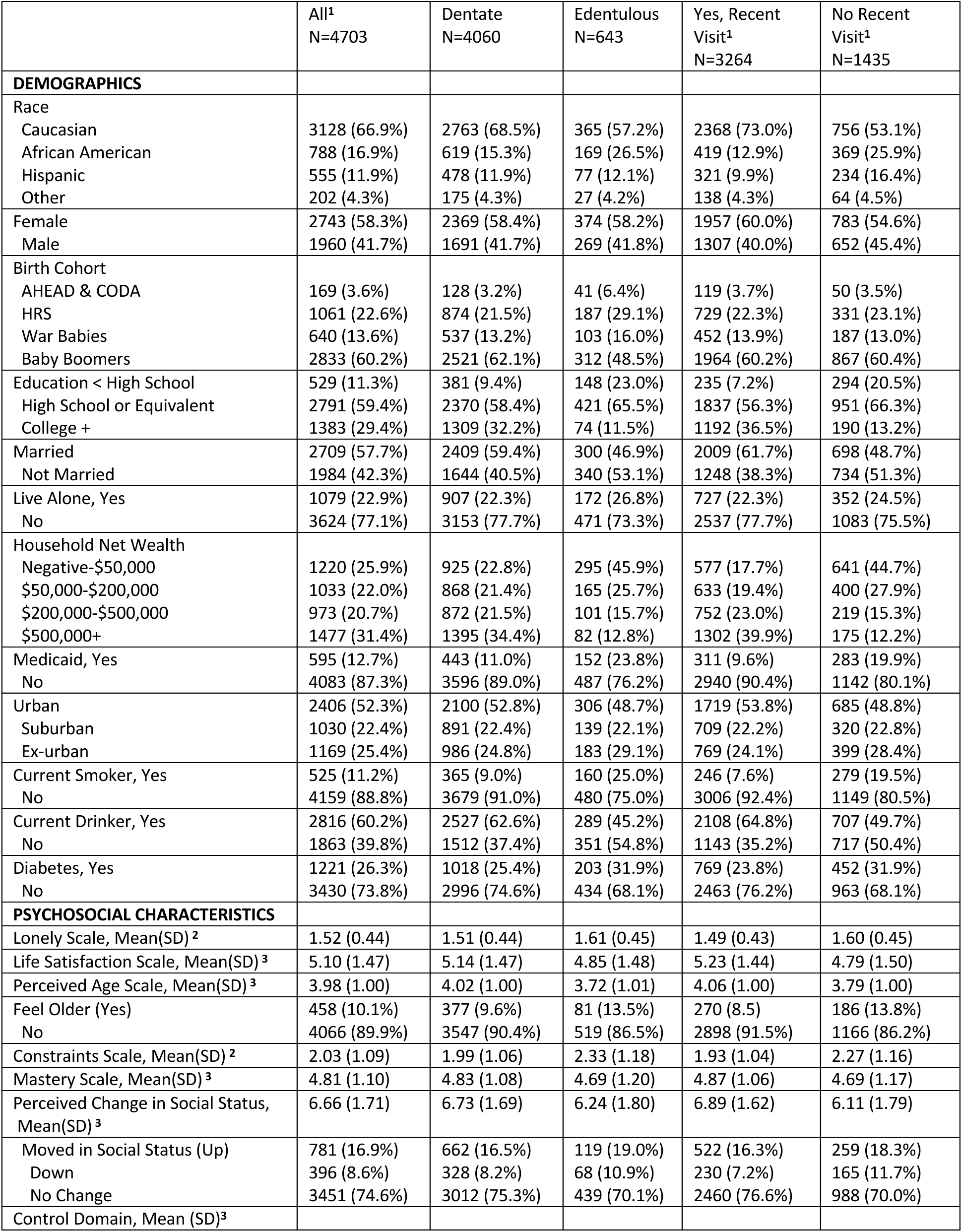
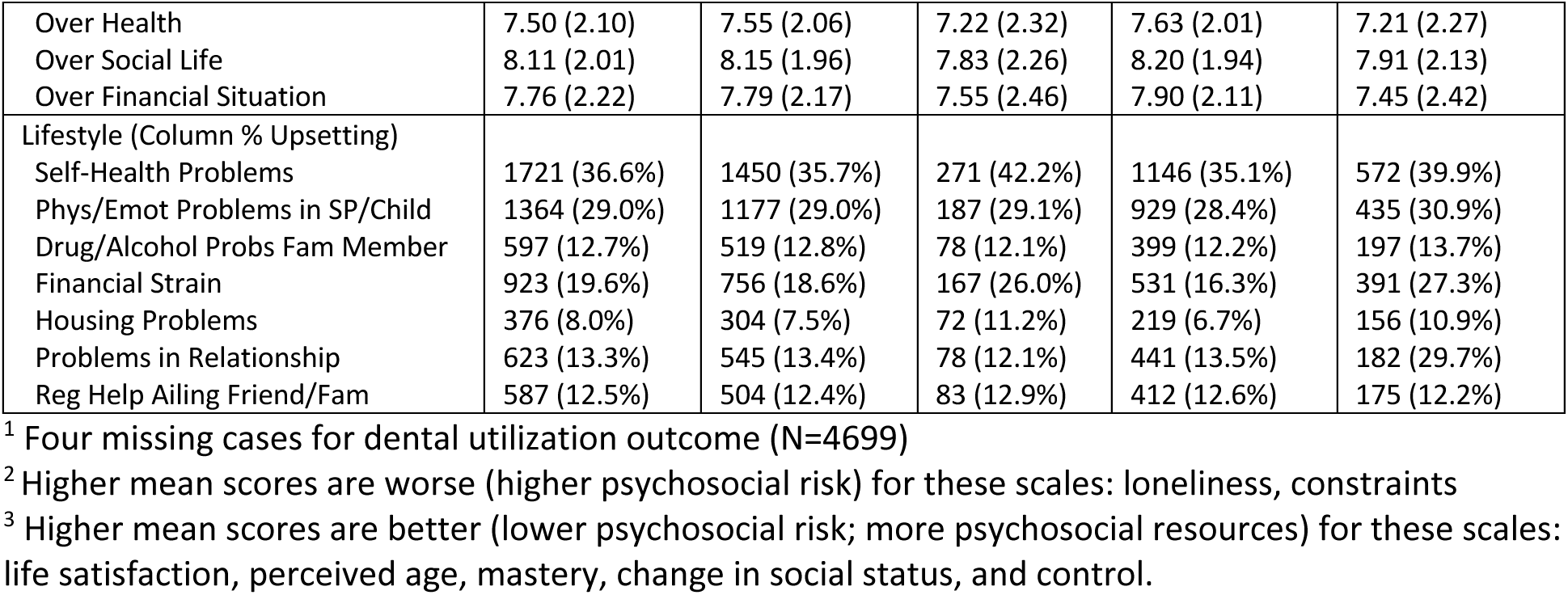
Demographic and Psychosocial Characteristics, by Dentition Status and Dental Visit in Last Two Years, 2018 (n=4703)

|  | All<sup>1</sup><br>N=4703 | Dentate<br>N=4060 | Edentulous<br>N=643 | Yes, Recent<br>Visit<sup>1</sup><br>N=3264 | No Recent<br>Visit<sup>1</sup><br>N=1435 |
| --- | --- | --- | --- | --- | --- |
| **DEMOGRAPHICS** |  |  |  |  |  |
| Race |  |  |  |  |  |
| Caucasian | 3128 (66.9%) | 2763 (68.5%) | 365 (57.2%) | 2368 (73.0%) | 756 (53.1%) |
| African American | 788 (16.9%) | 619 (15.3%) | 169 (26.5%) | 419 (12.9%) | 369 (25.9%) |
| Hispanic | 555 (11.9%) | 478 (11.9%) | 77 (12.1%) | 321 (9.9%) | 234 (16.4%) |
| Other | 202 (4.3%) | 175 (4.3%) | 27 (4.2%) | 138 (4.3%) | 64 (4.5%) |
| Female | 2743 (58.3%) | 2369 (58.4%) | 374 (58.2%) | 1957 (60.0%) | 783 (54.6%) |
| Male | 1960 (41.7%) | 1691 (41.7%) | 269 (41.8%) | 1307 (40.0%) | 652 (45.4%) |
| Birth Cohort |  |  |  |  |  |
| AHEAD & CODA | 169 (3.6%) | 128 (3.2%) | 41 (6.4%) | 119 (3.7%) | 50 (3.5%) |
| HRS | 1061 (22.6%) | 874 (21.5%) | 187 (29.1%) | 729 (22.3%) | 331 (23.1%) |
| War Babies | 640 (13.6%) | 537 (13.2%) | 103 (16.0%) | 452 (13.9%) | 187 (13.0%) |
| Baby Boomers | 2833 (60.2%) | 2521 (62.1%) | 312 (48.5%) | 1964 (60.2%) | 867 (60.4%) |
| Education < High School | 529 (11.3%) | 381 (9.4%) | 148 (23.0%) | 235 (7.2%) | 294 (20.5%) |
| High School or Equivalent | 2791 (59.4%) | 2370 (58.4%) | 421 (65.5%) | 1837 (56.3%) | 951 (66.3%) |
| College + | 1383 (29.4%) | 1309 (32.2%) | 74 (11.5%) | 1192 (36.5%) | 190 (13.2%) |
| Married | 2709 (57.7%) | 2409 (59.4%) | 300 (46.9%) | 2009 (61.7%) | 698 (48.7%) |
| Not Married | 1984 (42.3%) | 1644 (40.5%) | 340 (53.1%) | 1248 (38.3%) | 734 (51.3%) |
| Live Alone, Yes | 1079 (22.9%) | 907 (22.3%) | 172 (26.8%) | 727 (22.3%) | 352 (24.5%) |
| No | 3624 (77.1%) | 3153 (77.7%) | 471 (73.3%) | 2537 (77.7%) | 1083 (75.5%) |
| Household Net Wealth |  |  |  |  |  |
| Negative-\$50,000 | 1220 (25.9%) | 925 (22.8%) | 295 (45.9%) | 577 (17.7%) | 641 (44.7%) |
| \$50,000-\$200,000 | 1033 (22.0%) | 868 (21.4%) | 165 (25.7%) | 633 (19.4%) | 400 (27.9%) |
| \$200,000-\$500,000 | 973 (20.7%) | 872 (21.5%) | 101 (15.7%) | 752 (23.0%) | 219 (15.3%) |
| \$500,000+ | 1477 (31.4%) | 1395 (34.4%) | 82 (12.8%) | 1302 (39.9%) | 175 (12.2%) |
| Medicaid, Yes | 595 (12.7%) | 443 (11.0%) | 152 (23.8%) | 311 (9.6%) | 283 (19.9%) |
| No | 4083 (87.3%) | 3596 (89.0%) | 487 (76.2%) | 2940 (90.4%) | 1142 (80.1%) |
| Urban | 2406 (52.3%) | 2100 (52.8%) | 306 (48.7%) | 1719 (53.8%) | 685 (48.8%) |
| Suburban | 1030 (22.4%) | 891 (22.4%) | 139 (22.1%) | 709 (22.2%) | 320 (22.8%) |
| Ex-urban | 1169 (25.4%) | 986 (24.8%) | 183 (29.1%) | 769 (24.1%) | 399 (28.4%) |
| Current Smoker, Yes | 525 (11.2%) | 365 (9.0%) | 160 (25.0%) | 246 (7.6%) | 279 (19.5%) |
| No | 4159 (88.8%) | 3679 (91.0%) | 480 (75.0%) | 3006 (92.4%) | 1149 (80.5%) |
| Current Drinker, Yes | 2816 (60.2%) | 2527 (62.6%) | 289 (45.2%) | 2108 (64.8%) | 707 (49.7%) |
| No | 1863 (39.8%) | 1512 (37.4%) | 351 (54.8%) | 1143 (35.2%) | 717 (50.4%) |
| Diabetes, Yes | 1221 (26.3%) | 1018 (25.4%) | 203 (31.9%) | 769 (23.8%) | 452 (31.9%) |
| No | 3430 (73.8%) | 2996 (74.6%) | 434 (68.1%) | 2463 (76.2%) | 963 (68.1%) |
| **PSYCHOSOCIAL CHARACTERISTICS** |  |  |  |  |  |
| Lonely Scale, Mean(SD) <sup>2</sup> | 1.52 (0.44) | 1.51 (0.44) | 1.61 (0.45) | 1.49 (0.43) | 1.60 (0.45) |
| Life Satisfaction Scale, Mean(SD) <sup>3</sup> | 5.10 (1.47) | 5.14 (1.47) | 4.85 (1.48) | 5.23 (1.44) | 4.79 (1.50) |
| Perceived Age Scale, Mean(SD) <sup>3</sup> | 3.98 (1.00) | 4.02 (1.00) | 3.72 (1.01) | 4.06 (1.00) | 3.79 (1.00) |
| Feel Older (Yes) | 458 (10.1%) | 377 (9.6%) | 81 (13.5%) | 270 (8.5) | 186 (13.8%) |
| No | 4066 (89.9%) | 3547 (90.4%) | 519 (86.5%) | 2898 (91.5%) | 1166 (86.2%) |
| Constraints Scale, Mean(SD) <sup>2</sup> | 2.03 (1.09) | 1.99 (1.06) | 2.33 (1.18) | 1.93 (1.04) | 2.27 (1.16) |
| Mastery Scale, Mean(SD) <sup>3</sup> | 4.81 (1.10) | 4.83 (1.08) | 4.69 (1.20) | 4.87 (1.06) | 4.69 (1.17) |
| Perceived Change in Social Status, Mean(SD) <sup>3</sup> | 6.66 (1.71) | 6.73 (1.69) | 6.24 (1.80) | 6.89 (1.62) | 6.11 (1.79) |
| Moved in Social Status (Up) | 781 (16.9%) | 662 (16.5%) | 119 (19.0%) | 522 (16.3%) | 259 (18.3%) |
| Down | 396 (8.6%) | 328 (8.2%) | 68 (10.9%) | 230 (7.2%) | 165 (11.7%) |
| No Change | 3451 (74.6%) | 3012 (75.3%) | 439 (70.1%) | 2460 (76.6%) | 988 (70.0%) |
| Control Domain, Mean (SD)<sup>3</sup> |  |  |  |  |  |
| Over Health | 7.50 (2.10) | 7.55 (2.06) | 7.22 (2.32) | 7.63 (2.01) | 7.21 (2.27) |
| Over Social Life | 8.11 (2.01) | 8.15 (1.96) | 7.83 (2.26) | 8.20 (1.94) | 7.91 (2.13) |
| Over Financial Situation | 7.76 (2.22) | 7.79 (2.17) | 7.55 (2.46) | 7.90 (2.11) | 7.45 (2.42) |
| Lifestyle (Column % Upsetting) |  |  |  |  |  |
| Self-Health Problems | 1721 (36.6%) | 1450 (35.7%) | 271 (42.2%) | 1146 (35.1%) | 572 (39.9%) |
| Phys/Emot Problems in SP/Child | 1364 (29.0%) | 1177 (29.0%) | 187 (29.1%) | 929 (28.4%) | 435 (30.9%) |
| Drug/Alcohol Probs Fam Member | 597 (12.7%) | 519 (12.8%) | 78 (12.1%) | 399 (12.2%) | 197 (13.7%) |
| Financial Strain | 923 (19.6%) | 756 (18.6%) | 167 (26.0%) | 531 (16.3%) | 391 (27.3%) |
| Housing Problems | 376 (8.0%) | 304 (7.5%) | 72 (11.2%) | 219 (6.7%) | 156 (10.9%) |
| Problems in Relationship | 623 (13.3%) | 545 (13.4%) | 78 (12.1%) | 441 (13.5%) | 182 (29.7%) |
| Reg Help Ailing Friend/Fam | 587 (12.5%) | 504 (12.4%) | 83 (12.9%) | 412 (12.6%) | 175 (12.2%) |

<sup>1</sup> Four missing cases for dental utilization outcome (N=4699)
<sup>2</sup> Higher mean scores are worse (higher psychosocial risk) for these scales: loneliness, constraints
<sup>3</sup> Higher mean scores are better (lower psychosocial risk; more psychosocial resources) for these scales: life satisfaction, perceived age, mastery, change in social status, and control.

All demographic characteristics except sex and nearly all psychosocial factors had statistically significant differences by dentition status (p<0.05). All scale scores reflected higher risk among the edentulous group. Edentulous participants reported being lonely more frequently, less satisfied with life, felt negatively about their perceived age, and feel older than their chronological age than dentate adults. Persons who were edentulous reported more constraints, lower mastery, lower perceived SES, and felt they moved down SES ladder in last two years than dentate. Edentulous participants have lower efficacy in all domains (health, social life, financial; lower scores than dentate) and report chronic stressors related to health, financial strain, and housing than dentate. No significant differences were observed by dentition status for these chronic stressors: problems with spouse/child, drug/alcohol problems with family, relationship problems, or helping ailing friend/family.

When comparing groups by whether or not the respondent had a recent dental visit, all demographic characteristics were statistically significant, except for birth cohort. Nearly all psychosocial scale scores reflected higher risk among persons reporting no recent dental visit. Adults with no recent dental visit reported more frequently being lonely (higher score), less satisfied with life (lower score), feel negatively about their perceived age (lower score), and feel older than their chronological age than adults who have gone to dentist in last two years. Further, adults with no recent dental visit have more constraints (higher score), lower mastery (lower score), perceive themselves as lower SES (lower score), feel they have moved down SES ladder in last two years and have lower efficacy in all domains (health, social life, financial; lower scores than dentate). Adults with no recent dental visit report more chronic stressors related to health, financial strain, and housing than their counterparts. However, there were no statistically significant differences by dental visit for problems with spouse/child, drug/alcohol problems with family, relationship problems, or helping ailing friend/family.

**Table 3** summarizes the distribution of characteristics for each of the three LCA psychosocial classes for edentulism and no recent dental visit. About half the sample (n=2230) were in Class A:“Satisfied/Connected”, and about quarter were in Class B:“Satisfied/Lonely” (n=1293) and Class C:“Unsatisfied/Lonely” (n=1180), respectively. Class A consistently had the fewest psychosocial stressors and most resources. Class B was mixed, with scores generally between the other two classes; notably their psychosocial risk profile is closer to Class C on two constructs, loneliness and perceived negative impact of self-aging. Class C had the most psychosocial risk factors and fewest resources. Consistent patterns emerged. The demographic composition across the classes revealed differences by race (more African Americans and fewer Whites in Class C), sex (more females in Classes A and C), birth cohort (more Baby Boomers in Class C), marital status (more married in Class A), more on Medicaid in Class C. Persons in Classes A and B had greater educational attainment and household wealth than those in Class C. Class C had more smokers and diabetics.

**Table 3:**
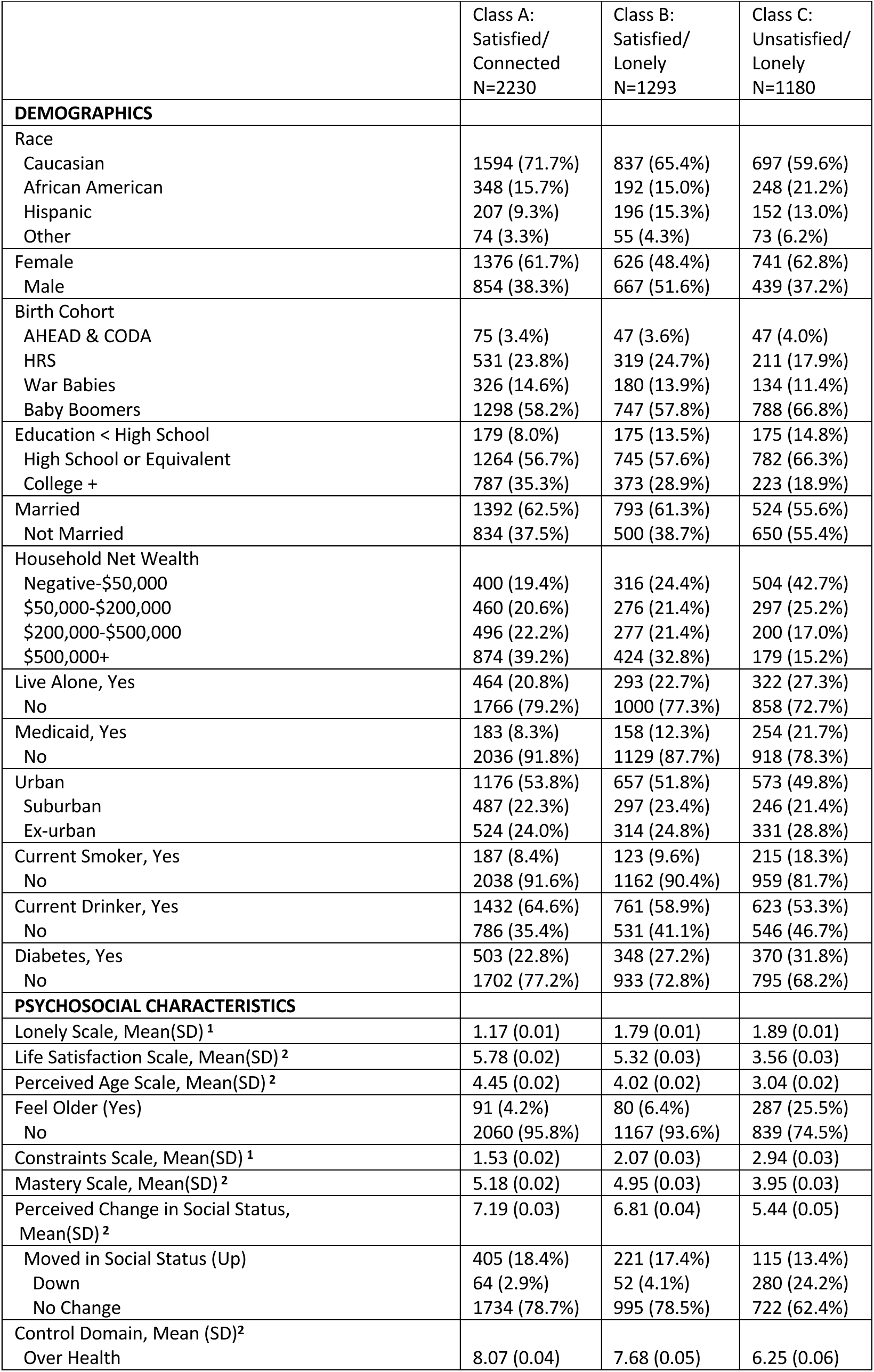
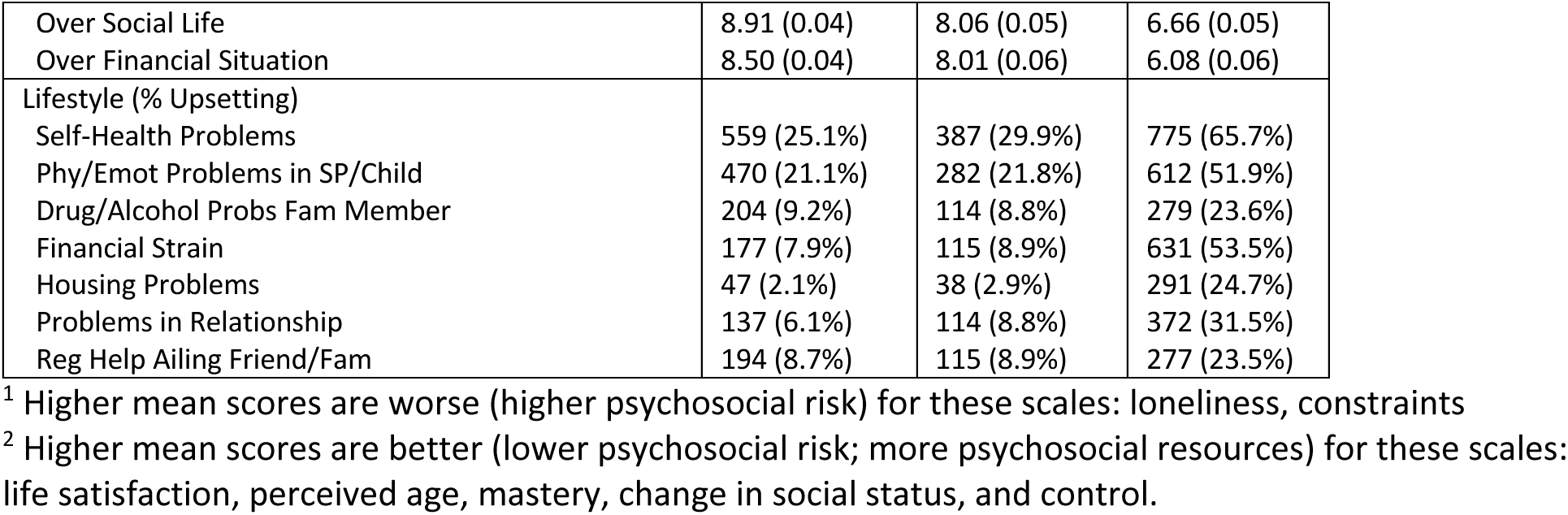
Demographic and Psychosocial Characteristics, by LCA Class, 2018 (n=4703)

|  | Class A:<br>Satisfied/<br>Connected<br>N=2230 | Class B:<br>Satisfied/<br>Lonely<br>N=1293 | Class C:<br>Unsatisfied/<br>Lonely<br>N=1180 |
| --- | --- | --- | --- |
| **DEMOGRAPHICS** |  |  |  |
| Race |  |  |  |
| Caucasian | 1594 (71.7%) | 837 (65.4%) | 697 (59.6%) |
| African American | 348 (15.7%) | 192 (15.0%) | 248 (21.2%) |
| Hispanic | 207 (9.3%) | 196 (15.3%) | 152 (13.0%) |
| Other | 74 (3.3%) | 55 (4.3%) | 73 (6.2%) |
| Female | 1376 (61.7%) | 626 (48.4%) | 741 (62.8%) |
| Male | 854 (38.3%) | 667 (51.6%) | 439 (37.2%) |
| Birth Cohort |  |  |  |
| AHEAD & CODA | 75 (3.4%) | 47 (3.6%) | 47 (4.0%) |
| HRS | 531 (23.8%) | 319 (24.7%) | 211 (17.9%) |
| War Babies | 326 (14.6%) | 180 (13.9%) | 134 (11.4%) |
| Baby Boomers | 1298 (58.2%) | 747 (57.8%) | 788 (66.8%) |
| Education < High School | 179 (8.0%) | 175 (13.5%) | 175 (14.8%) |
| High School or Equivalent | 1264 (56.7%) | 745 (57.6%) | 782 (66.3%) |
| College + | 787 (35.3%) | 373 (28.9%) | 223 (18.9%) |
| Married | 1392 (62.5%) | 793 (61.3%) | 524 (55.6%) |
| Not Married | 834 (37.5%) | 500 (38.7%) | 650 (55.4%) |
| Household Net Wealth |  |  |  |
| Negative-\$50,000 | 400 (19.4%) | 316 (24.4%) | 504 (42.7%) |
| \$50,000-\$200,000 | 460 (20.6%) | 276 (21.4%) | 297 (25.2%) |
| \$200,000-\$500,000 | 496 (22.2%) | 277 (21.4%) | 200 (17.0%) |
| \$500,000+ | 874 (39.2%) | 424 (32.8%) | 179 (15.2%) |
| Live Alone, Yes | 464 (20.8%) | 293 (22.7%) | 322 (27.3%) |
| No | 1766 (79.2%) | 1000 (77.3%) | 858 (72.7%) |
| Medicaid, Yes | 183 (8.3%) | 158 (12.3%) | 254 (21.7%) |
| No | 2036 (91.8%) | 1129 (87.7%) | 918 (78.3%) |
| Urban | 1176 (53.8%) | 657 (51.8%) | 573 (49.8%) |
| Suburban | 487 (22.3%) | 297 (23.4%) | 246 (21.4%) |
| Ex-urban | 524 (24.0%) | 314 (24.8%) | 331 (28.8%) |
| Current Smoker, Yes | 187 (8.4%) | 123 (9.6%) | 215 (18.3%) |
| No | 2038 (91.6%) | 1162 (90.4%) | 959 (81.7%) |
| Current Drinker, Yes | 1432 (64.6%) | 761 (58.9%) | 623 (53.3%) |
| No | 786 (35.4%) | 531 (41.1%) | 546 (46.7%) |
| Diabetes, Yes | 503 (22.8%) | 348 (27.2%) | 370 (31.8%) |
| No | 1702 (77.2%) | 933 (72.8%) | 795 (68.2%) |
| **PSYCHOSOCIAL CHARACTERISTICS** |  |  |  |
| Lonely Scale, Mean(SD) <sup>1</sup> | 1.17 (0.01) | 1.79 (0.01) | 1.89 (0.01) |
| Life Satisfaction Scale, Mean(SD) <sup>2</sup> | 5.78 (0.02) | 5.32 (0.03) | 3.56 (0.03) |
| Perceived Age Scale, Mean(SD) <sup>2</sup> | 4.45 (0.02) | 4.02 (0.02) | 3.04 (0.02) |
| Feel Older (Yes) | 91 (4.2%) | 80 (6.4%) | 287 (25.5%) |
| No | 2060 (95.8%) | 1167 (93.6%) | 839 (74.5%) |
| Constraints Scale, Mean(SD) <sup>1</sup> | 1.53 (0.02) | 2.07 (0.03) | 2.94 (0.03) |
| Mastery Scale, Mean(SD) <sup>2</sup> | 5.18 (0.02) | 4.95 (0.03) | 3.95 (0.03) |
| Perceived Change in Social Status,<br>Mean(SD) <sup>2</sup> | 7.19 (0.03) | 6.81 (0.04) | 5.44 (0.05) |
| Moved in Social Status (Up) | 405 (18.4%) | 221 (17.4%) | 115 (13.4%) |
| Down | 64 (2.9%) | 52 (4.1%) | 280 (24.2%) |
| No Change | 1734 (78.7%) | 995 (78.5%) | 722 (62.4%) |
| Control Domain, Mean (SD) <sup>2</sup><br>Over Health | 8.07 (0.04) | 7.68 (0.05) | 6.25 (0.06) |
| Over Social Life | 8.91 (0.04) | 8.06 (0.05) | 6.66 (0.05) |
| Over Financial Situation | 8.50 (0.04) | 8.01 (0.06) | 6.08 (0.06) |
| Lifestyle (% Upsetting) |  |  |  |
| Self-Health Problems | 559 (25.1%) | 387 (29.9%) | 775 (65.7%) |
| Phy/Emot Problems in SP/Child | 470 (21.1%) | 282 (21.8%) | 612 (51.9%) |
| Drug/Alcohol Probs Fam Member | 204 (9.2%) | 114 (8.8%) | 279 (23.6%) |
| Financial Strain | 177 (7.9%) | 115 (8.9%) | 631 (53.5%) |
| Housing Problems | 47 (2.1%) | 38 (2.9%) | 291 (24.7%) |
| Problems in Relationship | 137 (6.1%) | 114 (8.8%) | 372 (31.5%) |
| Reg Help Ailing Friend/Fam | 194 (8.7%) | 115 (8.9%) | 277 (23.5%) |

<sup>1</sup> Higher mean scores are worse (higher psychosocial risk) for these scales: loneliness, constraints
<sup>2</sup> Higher mean scores are better (lower psychosocial risk; more psychosocial resources) for these scales: life satisfaction, perceived age, mastery, change in social status, and control.

In terms of psychosocial characteristics, Class C exhibited the highest loneliness score, lowest life satisfaction score, and felt worse about aging. One-quarter felt older than their age, they were highest on constraints and lowest on mastery, about one-quarter moved down in social status, they had the lowest control scores for all domains (health, social life, finance), and reported substantially more chronic stressors (24%-66%). Class A older adults had the fewest psychosocial risk factors and most resources; Class B’s profile was between Classes A and C.

**Table 4** shows the minimally and fully adjusted odds ratios for the two dichotomous outcomes comparing LCA classes. In fully adjusted models, Class B older adults had 1.29 (95% CI:1.03-1.61) times greater odds than Class A to be edentulous, and 1.27 (1.07-1.50) time greater odds to not have a recent dental visit. Similarly, Class C older adults had 1.20 (0.96-1.51) times greater odds than Class A to be edentulous, and 1.33 (1.11-1.58) times greater odds to not have a recent dental visit. Odds ratio estimates were attenuated in the fully adjusted models over the minimally adjusted models, but overall, older adults in both Classes B and C remained more likely to experience poor oral health outcomes than Class A.

**Table 4:** Adjusted Estimates (CI) for Comparison of Dental Outcomes by LCA Class, 2018.

|  | Minimally Adjusted<sup>1</sup> |  |  | Fully Adjusted<sup>2</sup> |  |  |
| --- | --- | --- | --- | --- | --- | --- |
|  | Class A:<br>Satisfied/<br>Connected | Class B:<br>Satisfied/<br>Lonely | Class C:<br>Unsatisfied/<br>Lonely | Class A:<br>Satisfied/<br>Connected | Class B:<br>Satisfied/<br>Lonely | Class C:<br>Unsatisfied/<br>Lonely |
|  | Odds Ratio (95% CI) |  |  | Odds Ratio (95% CI) |  |  |
| Edentulism<sup>3</sup> | Ref | 1.56 (1.27-1.93) | 1.98 (1.61-2.43) | Ref | 1.29 (1.03-1.61) | 1.20 (0.96-1.51) |
| No Recent Dental Utilization<sup>3</sup> | Ref | 1.44 (1.23-1.69) | 2.06 (1.76-2.41) | Ref | 1.27 (1.07-1.50) | 1.33 (1.11-1.58) |

<sup>1</sup> Minimally Adjusted by Race, Sex and Birth Cohort
<sup>2</sup> Fully Adjusted by Race, Sex, Birth Cohort, Education, Marital Status, Wealth, Medicaid, Urban, Smoker, Alcohol Drinker, and Diabetes
<sup>3</sup> Covariate-adjusted logistic regression odds ratio estimates are based on the HRS-LB sample (N=4703)

## DISCUSSION

This study examined how psychosocial characteristics were related to edentulism and dental utilization in HRS. Results demonstrate important relationships between adverse psychosocial factors and poor oral health outcomes for older adults. LCA patterns were consistent; more Class C:“Unsatisfied/Lonely” older adults were edentulous and lacked routine dental visits than other older adults. Class C fared worse than Class B, who were also lonely and had poor perceptions of aging. However, Class B had more psychosocial resources and higher life satisfaction than Class C, and appeared to be more resilient. Class A had the lowest risk and most psychosocial resources and experienced the best outcomes.

A major risk factor in the well-being domain was loneliness. Loneliness is not identical to social isolation, though often used interchangeably.^26^ Together they have been described as an important mortality risk factor.^27,28^ Social isolation usually reflects being alone, and aspects of one’s social network and frequency of interacting with and connection to others. Feeling lonely depends on an individual’s emotional interpretation to being alone. Being physically alone does not necessarily mean an older adult has no social connections, or feels lonely. Crowe^29^ studied loneliness longitudinally in HRS between 2006-2014, and found that feeling persistently lonely increased risk of morbidity, advanced signs of aging, and death. Class C was lonely, with low life satisfaction. While some interventions could be delivered to engage Class C older adults to reduce social isolation, the emotional loneliness response may be more difficult to overcome. A recent review of three pre-pandemic studies found video calls could help reduce social isolation and loneliness among older adults.^30^ Perissinotto^28^ suggests other practical strategies to measure and address loneliness and social isolation.

The beliefs domain included psychosocial perceptions of aging and resources. Some scales captured changes over time (worsening social status and functioning), with some items directly comparing to prior years. Reported downward shifts may accurately reflect real changes. Other beliefs and perceptions like sense of control/efficacy are potentially more malleable. Psychosocial resources remain understudied, though have potential to be improved to bolster resilience. Class C older adults had fewer resources of any kind (financial, psychosocial) to draw upon, relative to other Classes. Class C was overrepresented among lower SES categories and indicated low control.

A much higher proportion of Class C older adults suffered chronic stressors. Other studies have also found that low socio-economic position and exposure to stressors over time can negatively affect overall health and oral conditions into older adulthood.^31,32^ Exposure to chronic stressors are more challenging – there is a need to address more upstream social determinants and facilitate access to a range of resources to counter negative effects. Stressors like housing problems are not easily remedied. Other chronic stressors queried were outside of the control of the individual, and pertained to friends, family members, or other relationships, where different types of support or interventions are needed.

Results highlight the need to integrate behavioral and oral health care, and to assess older adults for these modifiable psychosocial risk and resource factors. Dental providers should screen for loneliness and other psychosocial factors, especially for edentulous patients or those who do not make routine dental visits frequently. Referrals to social workers/behavioral health professionals may be appropriate to address psychosocial risk factors and chronic stressors to ameliorate their negative impact. While private practitioners still predominate in dentistry, the field is moving towards more integrated health care service delivery. Primary care providers are beginning to conduct social needs assessments, using short screening tools like PRAPARE^33^ and ICD-10 Z-codes to document psychosocial needs.^34^ Dental providers practicing in federally-qualified health centers may be well-positioned to assess and refer for social needs. Health departments also often have information and referral resources available that dental providers could provide patients with identified needs.

There are some data limitations; all measures were self-reported, so responses may have been affected by social desirability and recall biases. Many psychosocial constructs assessed perceptions and beliefs, which are subjective. The cross-sectional dataset means causality cannot be determined. Further, we do not know whether or not edentulous individuals had well-fitting, functional, comfortable dentures. HRS does not include any clinically-assessed oral health outcomes. However, HRS data are from a national US sample, with diverse socio-demographics. This study leveraged the large, rich HRS-LB Subsample A, which appeared comparable to the full HRS. Study strengths include good sample size, and rich set of psychosocial risk and resource factors assessed through validated scales.

In sum, psychosocial factors and exposure to stressors matter for oral health. People who are edentulous and without recent dental visits were more likely to be “Unsatisfied/Lonely.”

## Acknowledgements/Grant Funding

The Health and Retirement Study data is sponsored by the National Institute on Aging (grand number U01AG009740) and is conducted by the University of Michigan. This analysis was conducted with financial support from the National Institute of Dental and Craniofacial Research, R03DE030161-01.

## Data Availability/Ethics statement

Publicly available data used in this analysis are online at the Health and Retirement Study website: https://hrs.isr.umich.edu/data-products. The University of North Carolina at Chapel Hill Office of Human Research Ethics determined this secondary data analysis did not require approval.

**Supplemental material 1. Distribution of psychosocial variables, HRS-LB 2018 (n=4703)**. see separate file.

