## Supplemental file 1 for "Psychosocial Profiles of Older Adults by Dentition Status and Dental Utilization History"

**Supplemental Materials File. Psychosocial Profiles Oral Health of Older Adults**
**Finlayson, Moss, Jones, Preisser, Weintraub**
**Health and Retirement Study Leave-Behind (HRS-LB) Subsample A data (2018)**

For the Latent Class Analysis (LCA), 53 item responses were dichotomized. Each item was dichotomized depending on the scale the item belonged to.

**Supplemental Tables 1-4 and Supplemental Figure 1** show the original variable distributions for the items in the psychosocial variables in these three domains from the HRS-LB sample for the LCA analysis (N=4703).

HRS-LB variable names can be found online in the 2018 codebook:
https://hrs.isr.umich.edu/sites/default/files/meta/2018/core/codebook/h18lb_ri.htm

Items that required reverse coding for the scale score to be tabulated are also denoted with an asterisk. Scale scoring information was from the 2006-2016 HRS-LB Codebook:
https://hrs.isr.umich.edu/sites/default/files/biblio/HRS%202006-2016%20SAQ%20Documentation_07.06.17_0.pdf
See **Supplemental Table 1** for information about the well-being domain psychosocial variables. The 11 items in the loneliness scale were dichotomized "Often", vs "some of the time" or "hardly ever or never". The 5 items in the subjective life satisfaction scale were dichotomized as "disagree" vs "agree" with the category "neither agree or disagree" were grouped with the "agree" category. The 7 items in the life satisfaction (domains) scale were grouped as "completely satisfied" or "very satisfied" vs all others.

See **Supplemental Tables 2 and 3** for information about the beliefs domain psychosocial variables. The society ladder was dichotomized < 7 vs 7+. The change in society ladder was moved down vs others. The 8 items in the age scale was split agree vs disagree. The 5 items in the control and 5 items in the mastery scales were dichotomized as agree vs disagree. The 3 items in the efficacy scales were dichotomized as < 7 vs 7+.

See **Supplemental Table 4** for information about the lifestyle domain psychosocial variables. The 7 items in the chronic lifestyle stressors were grouped as "no, didn't happen" or "yes, but not upsetting" vs "yes, somewhat upsetting" and "yes, very upsetting".

**Supplemental Table 1. Three well-being domain psychosocial variable items, response options, and dichotomies for loneliness, subjective life satisfaction, and domain-specific life satisfaction[1]**

| **Construct (# items)**<br>**Question Stem**<br>***Dichotomy***<br>**Response Options** | **Construct (# items)**<br>**Question Stem**<br>***Dichotomy***<br>**Response Options** | **Construct (# items)**<br>**Question Stem**<br>***Dichotomy***<br>**Response Options** |
|---|---|---|
| **Loneliness**<br>**(11 items)** | **Subjective Life Satisfaction**<br>**(5 items)** | **Subjective well-being,**<br>**Domain-specific (7 items)** |
| HOW MUCH OF THE TIME DO YOU FEEL…<br>1=often, 2=some of the time,<br>3=hardly ever or never<br>*Dichotomy: "Often" vs*<br>*"some of the time" or "hardly ever or never"* | Please say how much you agree or disagree with the following statements. 1=strongly disagree,<br>7=strongly agree<br>*Dichotomy: "disagree" vs*<br>*"agree" or "neither agree or disagree"* | Please think about your life and situation RIGHT NOW. HOW SATISFIED ARE YOU WITH...<br>1=completely satisfied to<br>5 = not at all satisfied.<br>*Dichotomy: "completely satisfied" or "very satisfied" vs all others* |
| lack companionship | In most ways my life is close to ideal. | condition of the place where you live (home, apt) |
| left out | The conditions of my life are excellent. | The city or town you live in |
| Isolated | I am satisfied with my life. | daily life and leisure activities |
| in tune with people | So far, I have gotten the important things I want in life. | Your family life |
| alone | If I could live my life again, I would change almost nothing. | Your present financial situation |
| people can talk to | | The total income of your household |
| people can turn to | | Your health |
| people understand you | | |
| people feel close to | | |
| part of group | | |
| in common | | |

[1]Notably, these are different questions and response options than the loneliness and life satisfaction single items in the HRS CORE; the HRS-LB scales were used for this analysis.

**Supplemental Table 2. Beliefs domain psychosocial variable items, response options, and dichotomies for self-perceived social status, age, and aging**

| **Construct (# items)**<br>**Question Stem**<br>***Dichotomy***<br>**Response Options** | **Construct (# items)**<br>**Question Stem**<br>***Dichotomy***<br>**Response Options** | **Construct (# items)**<br>**Question Stem**<br>***Dichotomy***<br>**Response Options** |
|---|---|---|
| **Society Ladder**<br>**(1 item)** | **Self-perceived age**<br>**(1 item)** | **Self-perception of aging**<br>**(8 items)** |
| Think of this ladder as representing where people stand in our society. At the top of the ladder are the people who are the best off - those who have the most money, most education, and best jobs. At the bottom are the people who are the worst off - who have the least money, least education, and the worst jobs or no jobs. The higher up you are on this ladder, the closer you are to the people at the very top and the lower you are, the closer you are to the people at the very bottom. Please mark an X on the rung on the ladder where you would place yourself. Range: 1 (low) – 10 (high)<br>*Dichotomy: <7 vs 7+* | Has your position on the ladder changed within the last two years?<br>1=Yes moved up, 2=Yes moved down, 3=No<br>*Dichotomy: "Yes moved down" vs "Yes moved up" or "No"* | The next statements are about the way people feel about their age and about the things that happen as they get older. Please tell us how much you agree or disagree with each statement for you personally.<br>1=strongly disagree, 6=strongly agree<br>*Dichotomy: disagree vs agree* |
| 10 (high) | Yes moved up | Things keep getting worse as I get older |
| 9 | Yes moved down | I have as much pep as I did last year |
| 8 | No | The older I get, the more useless I feel. |
| 7 | | I am as happy now as I was when I was younger. |
| 6 | | As I get older, things are better than I thought they would be. |
| 5 | | So far, I am satisfied with the way that I am aging. |
| 4 | | The older I get, the more I have had to stop doing things that I liked. |
| 3 | | Getting older has brought with it many things that I do not like. |
| 2 | | |
| 1 (low) | | |

**Supplemental Table 3. Beliefs domain (continued) psychosocial variable items, response options, and dichotomies for sense of control**

| **Construct (# items)**<br>**Question Stem**<br>***Dichotomy***<br>**Response Options** | **Construct (# items)**<br>**Question Stem**<br>***Dichotomy***<br>**Response Options** | **Construct (# items)**<br>**Question Stem**<br>***Dichotomy***<br>**Response Options** |
|---|---|---|
| **Sense of Control – General / Constraints (5 items)** | **Sense of Control – General / Mastery (5 items)** | **Sense of Control – Domain Specific / Efficacy (3 items)** |
| Please say how much you agree or disagree with each of the following statements.<br>1=strongly disagree, 6=strongly agree<br><br>*Dichotomy: disagree vs agree* | Please say how much you agree or disagree with each of the following statements.<br>1=strongly disagree, 6=strongly agree<br>*Dichotomy: disagree vs agree* | Using a 0 to 10 scale where 0 means "no control at all" and 10 means "very much control," how would you rate the amount of control you have over your health these days?<br>Range (11 point scale): 0-10<br>*Dichotomy: <7 vs 7+* |
| I often feel helpless in dealing with the problems of life. | I can do just about anything I really set my mind to. | CONTROL OVER HEALTH |
| Other people determine most of what I can and cannot do. | When I really want to do something, I usually find a way to succeed at it. | CONTROL OVER SOCIAL LIFE |
| What happens in my life is often beyond my control. | Whether or not I am able to get what I want is in my own hands. | CONTROL OVER FINANCIAL SITUATION |
| I have little control over the things that happen to me. | What happens to me in the future mostly depends on me. | |
| There is really no way I can solve the problems I have. | I can do the things that I want to do. | |

**Supplemental Table 4. Lifestyle domain psychosocial variable items, response options, and dichotomies for chronic lifestyle stressors**

| **Construct (# items)**<br>**Question Stem**<br>***Dichotomy***<br>**Response Options** |
|---|
| **Ongoing Chronic Lifestyle Stressors**<br>**(8 items)** |
| Please read the list below and indicate whether or not any of these are current and ongoing problems that have lasted twelve months or longer. If the problem is happening to you, indicate how upsetting it has been. Check the answer that is most like your current situation.<br>1=no, didn't happen; 2=yes, but not upsetting; 3=yes, somewhat upsetting; 4=yes, very upsetting<br><br>*Dichotomy: "no, didn't happen" or "yes, but not upsetting" vs "yes, somewhat upsetting" or "yes, very upsetting"* |
| SELF HEALTH PROBLEMS |
| PHYSICAL /EMOTIONAL PROBLEMS IN SPOUSE/CHILD |
| DRUG/ALCOHOL PROBLEMS FAMILY MEMBER |
| DIFFICULTIES AT WORK |
| FINANCIAL STRAIN |
| HOUSING PROBLEMS |
| PROBLEMS IN RELATIONSHIP |
| REGULARLY HELP AILING FRIEND/FAMILY |

**Supplemental Figure 1. Distribution of Psychosocial Study Variables, HRS-LB (2018)**

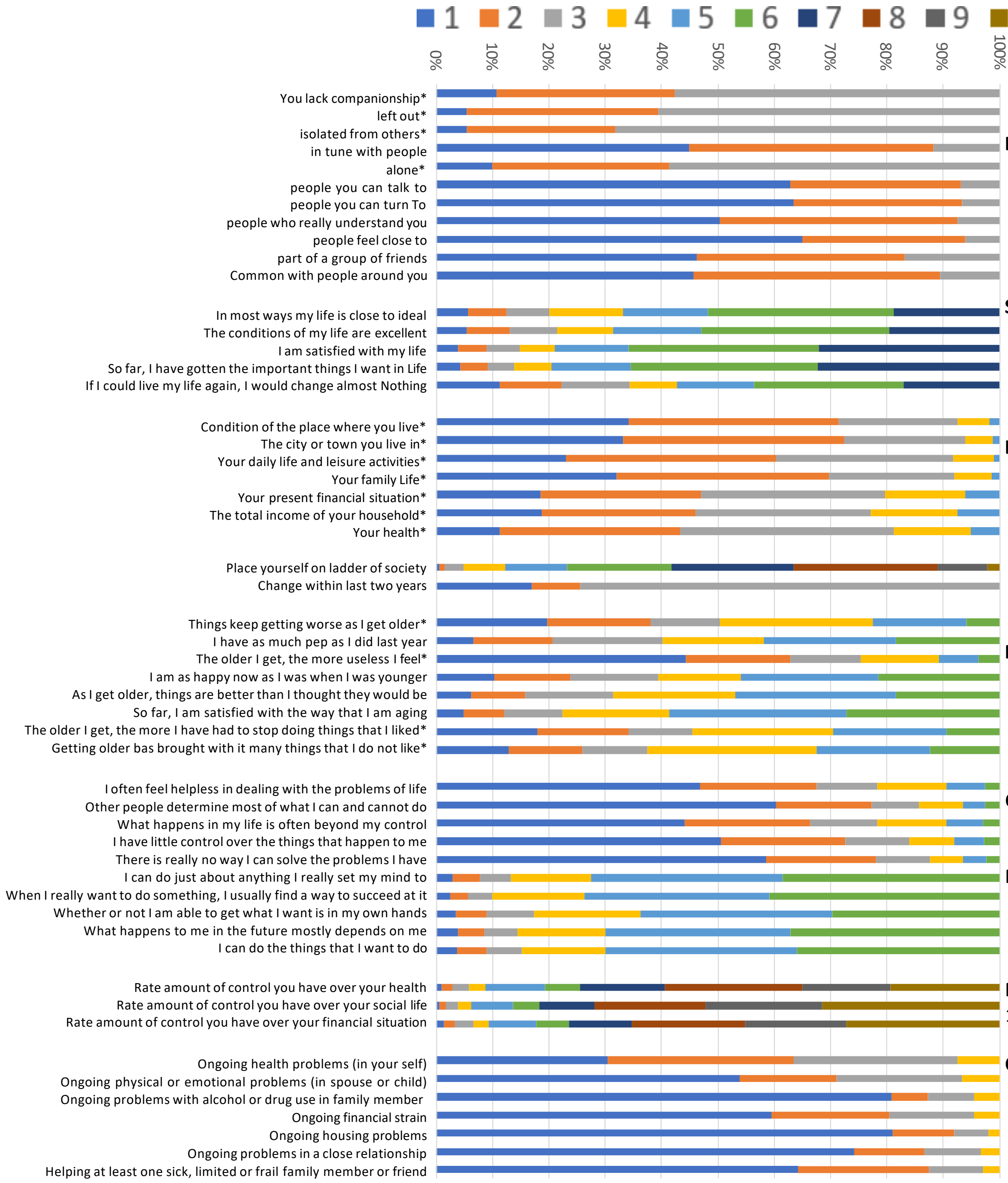


**Loneliness Scale “How much of the time do you feel…”**
1=Often
2=Some of the time
3=Hardly Ever or Never

**Subjective Life Satisfaction**
1=Strongly Disagree, 2=Somewhat Disagree, 3=Slightly Disagree
4=Neither Agree or Disagree, 5=Slightly Agree, 6=Somewhat Agree
7=Strongly Agree

**Life Satisfaction (Domain specific)**
1=Completely Satisfied, 2=Very Satisfied, 3=Somewhat Satisfied
4=Not Very Satisfied, 5=Not at All Satisfied

**Society Ladder:** 1 (low)-10 (High) Scale
Change within last two years 1=Moved Up, 2=Moved Down, 3=No Change

**Perceived Age**
1=Strongly Disagree, 2=Somewhat Disagree, 3=Slightly Disagree
4=Slightly Agree, 5=Somewhat Agree, 6=Strongly Agree

**Control**
1=Strongly Disagree, 2=Somewhat Disagree, 3=Slightly Disagree
4=Slightly Agree, 5=Somewhat Agree, 6=Strongly Agree

**Mastery**
1=Strongly Disagree, 2=Somewhat Disagree, 3=Slightly Disagree
4=Slightly Agree, 5=Somewhat Agree, 6=Strongly Agree

**Efficacy**
1 (low)-10 (High) Scale

**Chronic Lifestyle Stressors**
1=no, didn’t happen
2=yes, but not upsetting
3=yes, somewhat upsetting
4=yes, very upsetting

*Item Reversed in Scale
